# Metaproteogenomic Analysis of Saliva Samples from Parkinson’s Disease Patients Across the Spectrum of Cognitive Impairment

**DOI:** 10.1101/2022.12.29.22284030

**Authors:** Muzaffer Arıkan, Tuğçe Kahraman Demir, Zeynep Yıldız, Özkan Ufuk Nalbantoğlu, Nur Damla Korkmaz, Nesrin H. Yılmaz, Aysu Şen, Mutlu Özcan, Thilo Muth, Lütfü Hanoğlu, Süleyman Yıldırım

## Abstract

Cognitive impairment (CI) is very common in patients with Parkinson’s Disease (PD) and progressively develops on a spectrum from mild cognitive impairment (PD-MCI) to full dementia (PDD). Identification of PD patients at risk of developing cognitive decline, therefore, is unmet need in the clinic to manage the disease. Previous studies reported that oral microbiota of PD patients was altered even at early stages and poor oral hygiene is associated with dementia. However, data from single modalities often unable to explain complex chronic diseases in the brain and cannot reliably predict the risk of disease progression. Here, we performed integrative metaproteogenomic characterization of salivary microbiota and tested the hypothesis that biological molecules of saliva and saliva microbiota dynamically shift in association with the progression of cognitive decline and harbor discriminatory key signatures across the spectrum of CI in PD. We recruited a cohort of 115 participants in a multi-center study and employed multi-omics factor analysis (MOFA) to integrate amplicon sequencing and metaproteomic analysis to identify signature taxa and proteins in saliva. Our baseline analyses revealed contrasting interplay between the genus *Neisseria* and *Lactobacillus* and *Ligilactobacillus* genera across the spectrum of CI. The group specific signature profiles enabled us to identify candidate biomarkers including 7 bacterial genera (*Neisseria, Lactobacillus, Rothia, Ligilactobacillus, Alloprevotella, TM7x* and *Corynebacteirum*) and 4 protein groups (40S ribosomal protein SA, 40S ribosomal protein S15, pyruvate, phosphate dikinase and bactericidal permeability-increasing protein) discriminating CI stages in PD (AUC 0.74-0.86). Our study describes compositional dynamics of saliva across the spectrum of CI in PD and paves the way for developing novel, non-invasive biomarker strategies to predict the risk of CI progression in PD.

## 1. Introduction

Parkinson’s disease (PD) is a complex neurodegenerative disorder estimated to affect over 6 million people worldwide^1^. Due to the impact of the ageing population, a considerable increase in PD cases is expected in the future decades^2^. PD physiopathology is attributed to the alpha-synuclein (α-syn) aggregates accumulating in the neurons, which causes significant disruption of both motor and non-motor functions in the course of the disease^3^. One of its most common non-motor symptoms is cognitive impairment (CI) that progressively develops on a spectrum from mild cognitive impairment (MCI) to full-scale dementia (PDD)^4^. Despite variability among patients, there is a high risk of dementia in PD, with nearly half of patients reach the dementia stage within 10 years after diagnosis and virtually all patients develop full dementia within 20 years after diagnosis^5^. These patients cannot live independent lives and require care and support from their families and nursing homes, leading to economic burden. Thus, the current unmet need in the clinic is whether the PD patients at risk of developing cognitive decline can be predicted in order to implement disease changing interventions.

Saliva is a complex biofluid and considered to be a rich source of potential biomarkers for chronic diseases as saliva components typically include host cells, microbiota and biological molecules^6^. The oral health of PD patients such as saliva secretion, the composition of saliva, and dysbiosis significantly aggravate in the course of the disease^7^. Indeed, α-Syn can be detected in different biological fluids, including cerebrospinal fluid (CSF) and saliva^8–10^. The alpha-synuclein pathology in the oral cavity of PD patients often leads to poor secretion of saliva and dysphagia^11,12^. Remarkably, a six-year follow-up study of dysphagia in PD patients reported a significant association between CI and dysphagia^13^. Another common oral motor disorder of PD is drooling which has also been associated with CI in PD^14^. Together, these findings suggest a link between oral problems and CI in PD. Furthermore, recent studies reporting oral microbiota dysbiosis of PD patients linked dysphagia, drooling, and salivary pH with oral microbiota^15,16^. Therefore, we hypothesized that biological molecules of saliva and saliva microbiota dynamically shifts in association with CI progression in PD and harbors discriminatory key signature changes for predicting CI stages in PD.

In this study, using a comprehensive metaproteogenomics approach by combining 16S rRNA gene amplicon sequencing and metaproteomics, we recruited 115 subjects to identify changes in saliva composition that can be used to differentiate PD patients at different CI stages. We determined that salivary microbiota differentiates CI stages and detected bacterial taxa associated with CI. We also identified functional level changes associated with CI in PD. In addition, we integrated amplicon sequencing and metaproteomics data and identified a short list of candidate signatures differentiating CI stages in PD and evaluated their predictive performance through a machine learning model.

## 2. Materials and methods

### 2.1. Study subjects and clinical characteristics

The study was approved by the ethics committee of the Istanbul Medipol University with authorization number 10840098-604.01.01-E.3958, and informed consent was obtained from all participants. A total of 115 subjects (HC, n=27; PD-MCI, n=45; PDD, n=43) were recruited at two tertiary training hospitals including the Medipol Training and Research Hospital in the neurology clinic and Bakirkoy Research and Training Hospital for Psychiatric and Neurological Diseases. Participants were part of a larger cohort recruited into an ongoing prospective study on CI in PD. Clinical and demographic information, including age, sex, years of education were collected at clinic visits. The patients were examined by experienced neurologists and the diagnosis of PD was made within the framework of the “United Kingdom Parkinson’s Disease Society Brain Bank” criteria. Subjects with previous head trauma, stroke, or exposure to toxic substances, substance abuse, history of antibiotic or probiotic use within last one-month, chronic severe diseases (diabetes, cancer, kidney failure, etc.), autoimmune diseases, smokers, and those with symptoms suggestive of Parkinson’s plus syndromes were excluded from the study. Hoehn-Yahr Stages Parkinson’s Staging Scale was used to determine the stage of the disease and The Movement Disorder Society’s diagnostic criteria for Parkinson’s Disease Dementia criteria were used for dementia evaluation. The diagnosis of MCI was made within the framework of the criteria defined by Litvan et al.^17^

### 2.2. Sample preparation for 16S rRNA gene amplicon gene sequencing

Unstimulated saliva samples were divided into two equal aliquots of 500 µl and used for DNA and protein extractions separately. Microbial DNA extraction from saliva samples was performed using DNeasy PowerSoil (Qiagen, Hilden, Germany) with modifications as described before^18^. In brief, 250 µl saliva sample was centrifuged at 10,000 x g for 5 min, supernatant discarded, and the pellet was resuspended with 400 μl bead beating buffer and transferred to the PowerBead tube. Samples were homogenized by bead-beating using Next Advance Bullet Blender (30 s at level 4, 30 s incubation on ice and 30 s at level 4). After bead-beating step, the manufacturer’s protocol was followed without any modification. The V3-V4 regions of 16S rRNA gene were amplified using the universal bacterial primers (F-5′-CCTACGGGNGGCWGCAG-3′ and R-5′-GACTACHVGGGTATCTAATCC-3′). Next, amplicon libraries were prepared by following Illumina’s 16S rRNA metagenomic sequencing library preparation protocol and sequenced using a MiSeq platform and 2×250 paired end kit. Amplicon sequencing libraries prepared from a total of 115 gDNA samples were sequenced along with DNA extraction negative control and a no-template PCR control per run.

### 2.3. Sample preparation for metaproteomics

For protein extraction, 250 µl saliva sample was centrifuged at 10,000 x g for 5 min Discarding the supernatant, the pellet was resuspended in 250 μl bead beating buffer and transferred to the BeadBug™ prefilled tubes, 2.0 ml containing 1.0 mm Zirconium beads. Samples were homogenized by bead-beating using Next Advance Bullet Blender (30 s at level 4, 30 s incubation on ice and 30 s at level 4). After bead-beating step, samples were incubated at 100 °C for 10 min under constant shaking (600 rpm), followed by 4 °C incubation for an hour and centrifugation at 16,000 × g for 10 min. The supernatants were finally transferred to a clean 1.5 ml microcentrifuge tube. The total protein concentrations were measured with the Qubit protein assay kit. 50 µg protein (eluted in 30 µl) was used in the downstream analyses for each sample. Tryptic peptides were generated using the Filter Aided Sample Preparation Protocol (FASP) kit (Expedeon, San Diego, USA) according to the manufacturer’s protocol. The peptides were dissolved in 0.1 percent formic acid and diluted to 100 ng/μl before injecting to the liquid chromatography tandem-mass spectrometry (the ACQUITY UPLC M-Class coupled to a SYNAPT G2-Si high-definition mass spectrometer (Waters, Milford, CT)). The LC-MS/MS analysis was performed according to a previously published protocol^19^.

### 2.4. Analysis of 16S rRNA gene amplicon sequencing data

Raw 16S rRNA gene amplicon sequencing data were analyzed using the Nephele platform (v.1.6, http://nephele.niaid.nih.gov)^20^ using default parameters (QC: minlen: 30, req_qual: 12, overlap: 3, trail_qual: 3, run_flash2_merge: True, f2_min_overlap: 10, run_flash2_merge: True, error_rate: 0.1, window_size: 4; DADA2: maxEE: 5, ref_db: sv138.1, taxmethod: rdp, trimleft_fwd: 0, truncLen_fwd: 0, truncQ: 4) and SILVA v.138.1 database^21^. The contaminant sequences were identified and removed using the *decontam* package^22^ based on negative control samples. Only ASVs present in at least 2 samples were included in the downstream analyses. Samples were rarefied to minimum sampling depth (4,821 reads) before performing alpha diversity analyses and CLR transformed before beta diversity analyses. Diversity analyses were performed using QIIME2^23^ and phyloseq^24^. MaAsLin2^25^, LEfSe^26^, ANCOM-BC^27^ and ANOVA were used to determine differential abundances between groups, controlling covariates when possible, and the R package ggplot2^28^ was used for visualization of the results.

### 2.5. Analysis of metaproteomics data

For metaproteomics data analysis, firstly, a custom protein database based on 16S rRNA amplicon sequencing results was built. Briefly, we determined the 20 most abundant genera and bacterial taxa that showed differential abundance between study groups. We obtained all species-level genome bins that belonged to these genera from a recently published comprehensive study on human oral microbiome^29^. Next, we added all human proteins from UniProt database^30^ to the predicted bacterial proteins of these genome bins which produced a final protein database of 1,165,589 protein sequences. Progenesis-QI (Waters) software was used to identify and quantify the protein groups. Protein groups identified by at least 2 unique peptide sequences, taxonomically assigned to *Eukaryota* or Bacteria kingdoms and present in all samples were used for downstream analyses. Also, to reduce batch effects on metaproteomics results, we used MMUPHin^31^ to adjust protein abundances before further processing. Diversity analyses were performed using phyloseq. We employed Prophane^32^ tool with default parameters to annotate taxonomy and functions of the detected proteins. All identified protein groups from each sample were classified into main metabolic pathways using the orthologous groups (OG) classification against the EggNog database^33^. ANOVA was used to examine potential associations at both the protein OGs and functional categories levels with CI stages.

### 2.6. Multi-omics factor analysis

Multi-Omics Factor Analysis (MOFA)^34^ was employed to integrate 16S rRNA amplicon sequencing and metaproteomics datasets as data modalities with matching samples. ASV table was collapsed to the genus level and both datasets were CLR transformed to reduce compositionality bias before generating the MOFA model. Data and training options were set to default with 10,000 iterations in ‘slow’ convergence mode to generate 15 factors. Downstream analyses after generation of MOFA model were performed using MOFA+ tool^35^.

### 2.7. Model-based classification of CI status in PD

To identify taxonomic and protein signatures associated with CI, we constructed a random forest machine learning (ML) model. We first took advantage of the MOFA to select important features from the two omics datasets and subsequently fit these features into random forest models, both together and separately. To create the classifiers, a random forest constituted of 5000 trees was computed using microeco R package (v4.6-7)^36^. A total of 80% of samples were used to train the model and 20% were used for cross validation. We plotted the area under the receiver operating characteristic curve (AUROC) to show prediction performance of each model. To evaluate the predictive performance of our MOFA selected features, we have also used AutoGluon (v0.5.2) in tabular prediction mode. Evaluation metric ‘accuracy’ was chosen as the validation criteria. Bagging and stacking were enabled using “num_bag_folds=3”, “num_bag_sets=5”, “num_stack_levels=2” options. Testing was employed by external 10-fold cross-validation.

### 2.8. Statistical analyses

Statistical analyses were conducted in R 3.6.1. A Kruskal-Wallis test was used for alpha diversity comparisons. Adonis2, an implementation of permutational multivariate analysis of variance (PERMANOVA) from the vegan package with 999 permutations, was used for beta diversity comparisons, with adjustment for potential confounding factors. A t-test was used for continuous variables, namely age and education, while Fisher’s exact test was used for categorical variables. All P values, where appropriate, were adjusted for multiple testing using the Benjamini-Hochberg method.

### 2.9. Data availability

The raw 16S rRNA gene amplicon sequencing data produced in this study have been deposited in the NCBI Sequence Read Archive database under accession no. PRJNA913682. Metaproteomics data generated and analyzed in this study are available from the corresponding author on reasonable request.

## 3. Results

### 3.1. Characteristics of participants and analyses

A total of 115 individuals (43 PDD, 45 PD-MCI) and 27 HC were included in this study. Both 16S rRNA gene amplicon sequencing based microbiome analysis and metaproteomics profiling were performed for all salivary samples collected from the participants (Figure 1).

**Figure 1.**
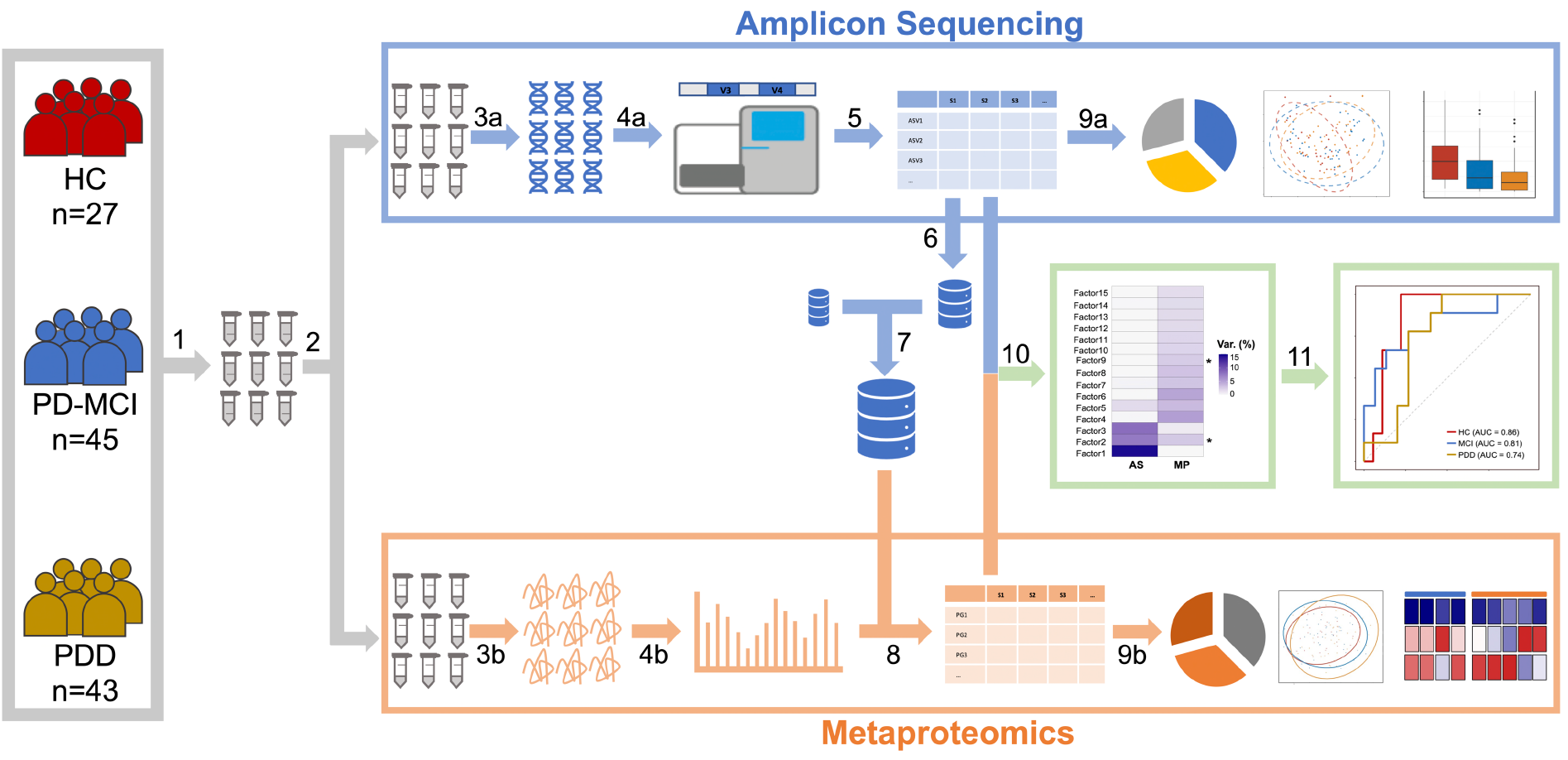
Experimental overview. (1) Saliva samples were collected from a total of 115 individuals (43 PDD, 45 PD-MCI) and 27 HC. (2) Samples were divided into two equal aliquots and used for DNA and protein extractions separately. (3a,b) DNA and protein isolations were performed. (4a,b) Amplicon libraries and tryptic peptides were prepared for NGS and LC-MS/MS, respectively. (5) ASV abundances were determined. (6) Predicted bacterial proteins of species-level genome bins that belonged to the 20 most abundant genera and bacterial taxa that showed differential abundance between study groups in amplicon sequencing were determined. (7) All human proteins from UniProt database were added to the predicted bacterial proteins which produced a final protein database that was used for protein identifications. (8) Protein group abundances were determined. (9a,b) Analyses of amplicon sequencing and metaproteomics data were performed. (10) Amplicon sequencing and metaproteomics datasets were integrated using MOFA. (11) MOFA selected important features from the two omics datasets were fit into random forest models to identify taxonomic and protein signatures associated with CI in PD.

The demographic and clinical features of the study participants are summarized in Table 1. The mean age differed significantly between the study groups. We therefore adjusted all P values for Age confounder, where appropriate. There was no significant difference in the proportion of female subjects between study groups. Years of education differed significantly between PD-MCI and HC, PDD and HC but not between PD-MCI and PDD participants which was also adjusted. There was expectedly a significant difference in mean MMSE scores between all study groups. The HC group had a mean MMSE score of 27.9, MCI group had a mean MMSE score of 23.6, and PDD group had a mean MMSE score of 18.7.

**Table 1.** Demographic and clinical features of the study cohort.

| Characteristics | HC | PD-MCI | PDD |
| --- | --- | --- | --- |
|  | (n=27) | (n=43) | (n=45) |
| Age (years, mean ± SD) | 59.6±8.20 | 67.1±8.48* | 71.4±7.10*<sup>#</sup> |
| Sex (Female) | 15 (55.6%) | 17 (39.5%) | 21 (46.7%) |
| Education (years, mean ± SD) | 10.8±5.1 | 7.4±4.8* | 4.6±4.5* |
| MMSE (mean ± SD) | 27.9±1.9 | 23.6±2.9* | 18.7±3.6*<sup>#</sup> |
| CDR (mean ± SD) | 0.0±0.0 | 0.5±0.0* | 1.2±0.5*<sup>#</sup> |
| Hoehn and Yahr score (mean ± SD) | - | 1.9±0.8 | 2.6±0.9<sup>#</sup> |
| UPDRS-part II (mean ± SD) | - | 32.3±14.9 | 50.0±16.8<sup>#</sup> |
| PD Duration (months) (mean ± SD) | - | 68.9±44.5 | 105.2±56.9<sup>#</sup> |

PD: Parkinson's Disease; SD: Standard Deviation; MMSE: Mini-Mental State Examination; HC: Healthy Control; PD-MCI: Parkinson's Disease with Mild Cognitive Impairment; PDD: Parkinson's Disease with Dementia
\* $p < 0.05$ for pairwise comparison with HC
<sup>#</sup> $p < 0.05$ for pairwise comparison with PD-MCI

### 3.2. Comparison of 16S rRNA based and metaproteomics based microbial composition results

We performed both 16S rRNA gene amplicon sequencing and metaproteomics for all 115 salivary samples. First, we compared ASV-based taxonomic composition with metaproteomics based taxonomic composition.

The 5 most abundant genera in salivary microbiota samples across sample groups were *Streptococcus, Prevotella, Veillonella, Campylobacter* and *Neisseria* in both 16S rRNA gene amplicon and metaproteomics results (Figure 2A). However, the relative abundance distributions of bacterial genera did not show any correlation between two methods which probably results from relatively higher/lower expression of proteins in some bacterial genera in saliva as expected (Figure 2B). We calculated genus, family, and phylum level overlap between the methods. The overlap at genus level was 12.4% which increased to 19.7% at family level and 46.2% at phylum level (Figure 2C-E). All shared genera, families and phyla are shown in Supplementary Figure 1. The amplicon sequencing detected much more bacterial taxa at all three levels than metaproteomics profiling. This is not surprising because only a subset of bacterial genera (as described in the Methods) were used to generate a custom-based reference protein database to identify proteins. In addition, we observed a larger fluctuation of bacterial relative abundances based on 16S rRNA gene amplicon sequencing across samples than that of metaproteomics.

**Figure 2.**
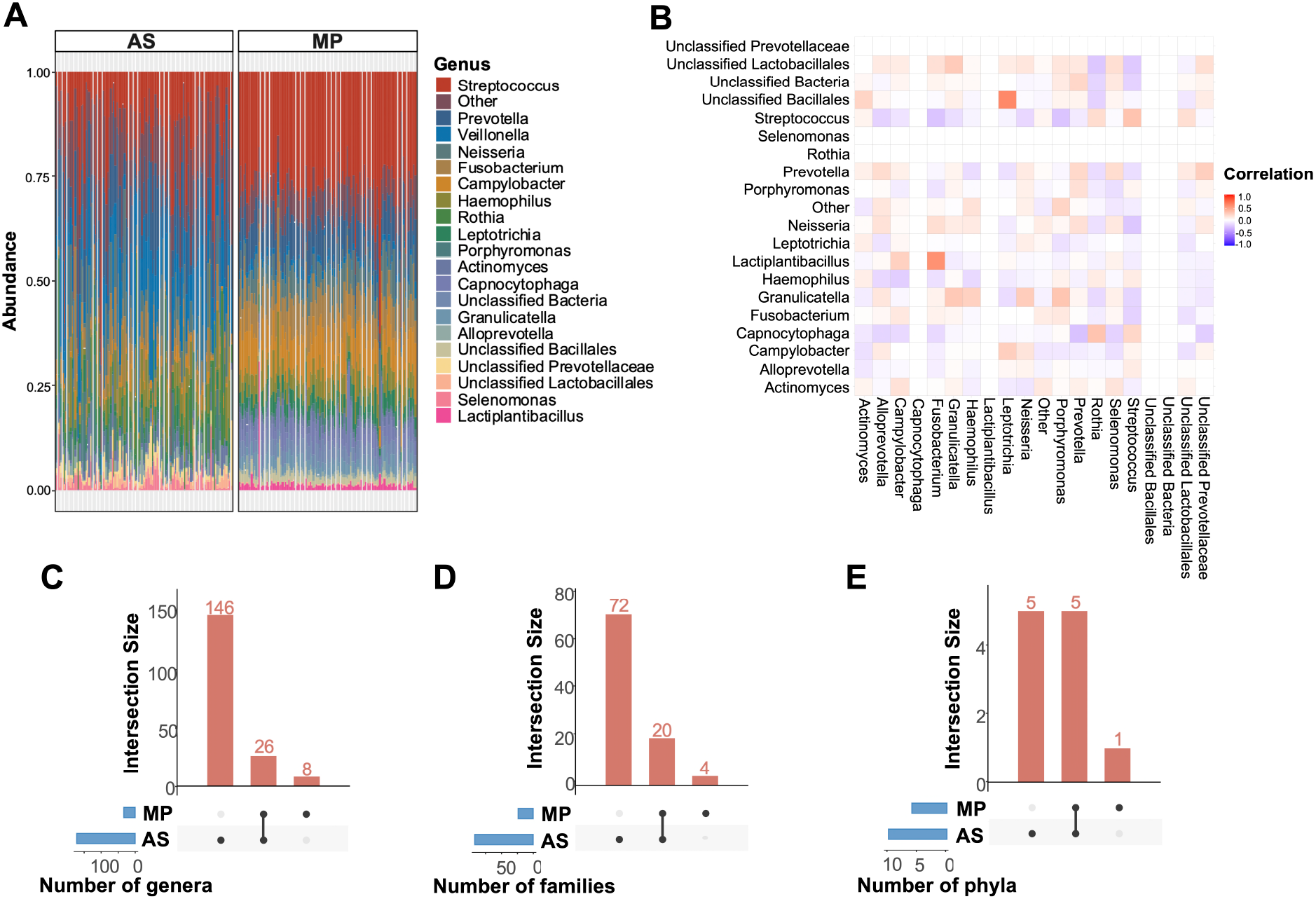
16S rRNA gene amplicon sequencing (AS) and metaproteomics (MP) based overview of salivary microbiota composition across samples. (A) The 20 most common bacterial genera in salivary microbiota samples by 16S rRNA gene amplicon sequencing and metaproteomics. Genera that were not among 20 most common taxa were grouped into “Other.” Each bar represents relative abundance distribution for a sample. The order of sample bars is same for both methods. (B) Correlation between abundance of bacterial genera by 16S rRNA gene amplicon sequencing versus metaproteomics. (C-E) Number of genera, families and phyla found across 16S rRNA gene amplicon sequencing and metaproteomics. Vertical bars represent the number of taxa shared between two methods highlighted with connected dots in the lower panel. Horizontal bars in the lower panel indicate the total number of taxa detected by each method.

### 3.3. 16S rRNA gene amplicon-based microbiome profiles differentiate study groups

We calculated alpha and beta diversity metrics for saliva samples based on 16S rRNA gene amplicon sequencing data. There were no significant differences in alpha diversity indices (Chao, Shannon, InvSimpson and Fisher) between study groups (Figure 3A). On the other hand, beta diversity analyses showed significant differences between study groups (Figure 3B). We generated a beta-diversity ordination using the Aitchison distance and tested if the samples cluster beyond expected by a chance while adjusting for the confounding effects of age, sex, and education. The results showed a significant difference between the study groups (PERMANOVA, R^2^ = 0.021, *p* = 0.024). To strengthen the conclusion, we also we used the Bray-Curtis and Jaccard distance for 16S rRNA gene amplicon sequencing results to test differences between study groups, which also showed a significant separation between the three groups (Bray-Curtis, PERMANOVA, R^2^ = 0.024, *p* = 0.021; Jaccard, PERMANOVA, R2 = 0.021, *p* = 0.017) (Supplementary Figure 2).

**Figure 3.**
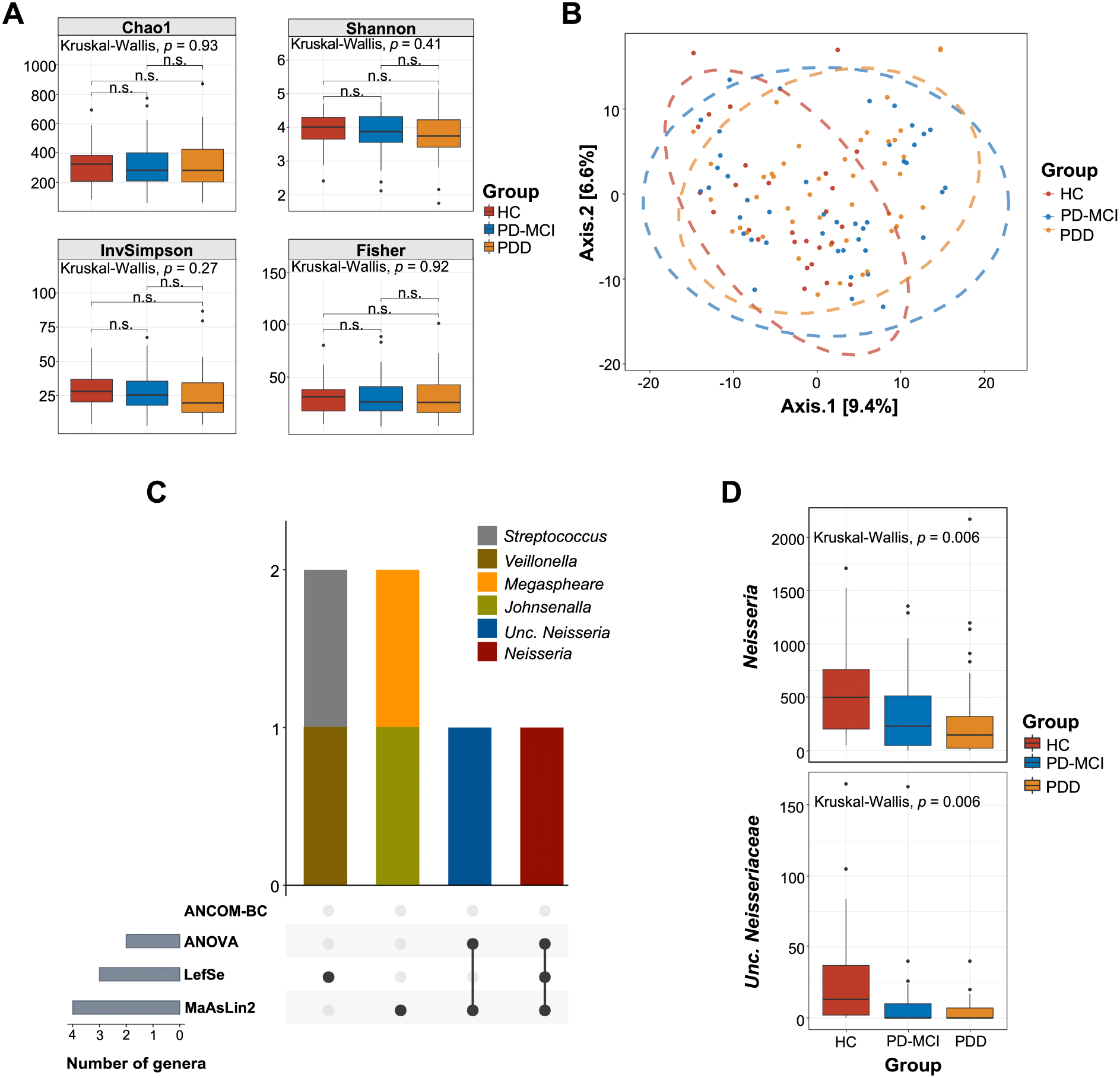
Structural diversity and differential abundance analysis of saliva samples by 16S rRNA gene amplicon sequencing. (A) Alpha diversity (Chao1, Shannon, InvSimpson, Fisher) comparisons of salivary microbiota samples between study groups. Median estimates compared across study groups using the Kruskal-Wallis test. Boxes represent the interquartile range, lines indicate medians, and whiskers indicate the range. n.s: not significant. (B) Beta diversity comparisons of saliva samples between study groups. PCoA was calculated using Aitchison distance. Ellipses represent an 95% confidence level. Color is indicative of the study group. (C) Number of differentially abundant genera across 4 microbiome differential abundance methods, namely ANCOM-BC, ANOVA, LefSe, MaAsLin2.Vertical bars represent the number of differentially abundant genera shared between the methods highlighted with connected dots in the lower panel. Horizontal bars in the lower panel indicate the total number of differentially abundant genera detected by each method. (D) Abundance distribution of differentially abundant genera detected by at least two methods across study groups. Median estimates compared across study groups using the Kruskal-Wallis test. Boxes represent the interquartile range, lines indicate medians, and whiskers indicate the range. *p* values represent the overall FDR-corrected *p* values.

To determine which microbial taxa were significantly associated with CI, we performed differential abundance using three different tools, namely LEfSe, MaAsLin2, ANCOM-BC and ANOVA (Figure 3C). Because the correct identification of differentially abundant microbial taxa between experimental conditions vary between different methods^37^, we only reported taxa determined by a minimum of two different methods in consensus. ANOVA-based differential abundance analysis showed a significant decrease in the abundance of *Neisseria* genus and unclassified *Neisseriacceae* with the progression of CI (Figure D) while LEfSe-based analysis not only showed a decrease in *Neisseria* genus but also an accompanied significant increase in the abundance of *Streptococcus* and *Veillonella* genera with the progression of CI. The decrease in the abundance of *Neisseria* was further supported by MaAsLin2 analysis while ANCOM-BC did not detect any significant difference between study groups. We further investigated potential correlations among bacterial genera and with the covariates MMSE, CDR, UPDRSII. We determined a significant positive correlation between *Neisseria* and MMSE (p=0.003, Spearman’s ρ=0.268) and a significant negative correlation between *Neisseria* and CDR score (p=0.004, Spearman’s ρ=-0.27) (Supplementary Figure 3). We have also detected a negative correlation of *Lactobacillus* and *Ligilactobacillus* with *Neisseria* (p=0.003, Spearman’s ρ=-0.27 and p=0.03, Spearman’s ρ=-0.20, respectively) (Supplementary Figure 3).

### 3.4. Metaproteomic profiling identifies human and microbial proteins in saliva

We applied metaproteomics to determine the functional characteristics of the saliva and quantified a total of 29,054 unique peptides and 9,379 protein groups in 115 samples. Filtering protein groups that were not identified by at least 2 unique peptide sequences yielded 4253 proteins across our samples. Then, we removed protein groups that were not taxonomically assigned as *Eukaryota* or *Bacteria* which reduced the number of proteins to 3354. Finally, proteins that were not detected in all samples were filtered out to control the batch effect and the most robustly quantified 537 proteins were used for downstream analyses.

Among 537 protein groups quantified, 287 (53.5%) were from salivary microorganisms while 250 (46.5%) protein groups were from human proteome origin. Human protein groups constituted 67.9% of the total protein intensities measured in the salivary samples while microbial protein groups constituted 32.1% of total protein intensities (Figure 4A) which shows that human proteins are more abundant in saliva and consistent with previous studies^38^. We tested if the ratio between human and microbial protein groups changes with the progression of the CI and detected no significant change (Supplementary Figure 4). We also performed beta diversity analysis based on human and microbial proteins separately to evaluate if there is a significant association between human or microbial protein group composition and CI. The results did not show any compositional difference in only microbial or only human protein groups profiles across study groups (Figure 4B and C). However, beta diversity analysis based on all detected proteins showed significant separation between groups (PERMANOVA, R2 = 0.026, p = 0.025); although when adjusted for all potential confounders, the difference was attenuated (PERMANOVA, R2 = 0.019, p = 0.082) (Figure 4D).

**Figure 4.**
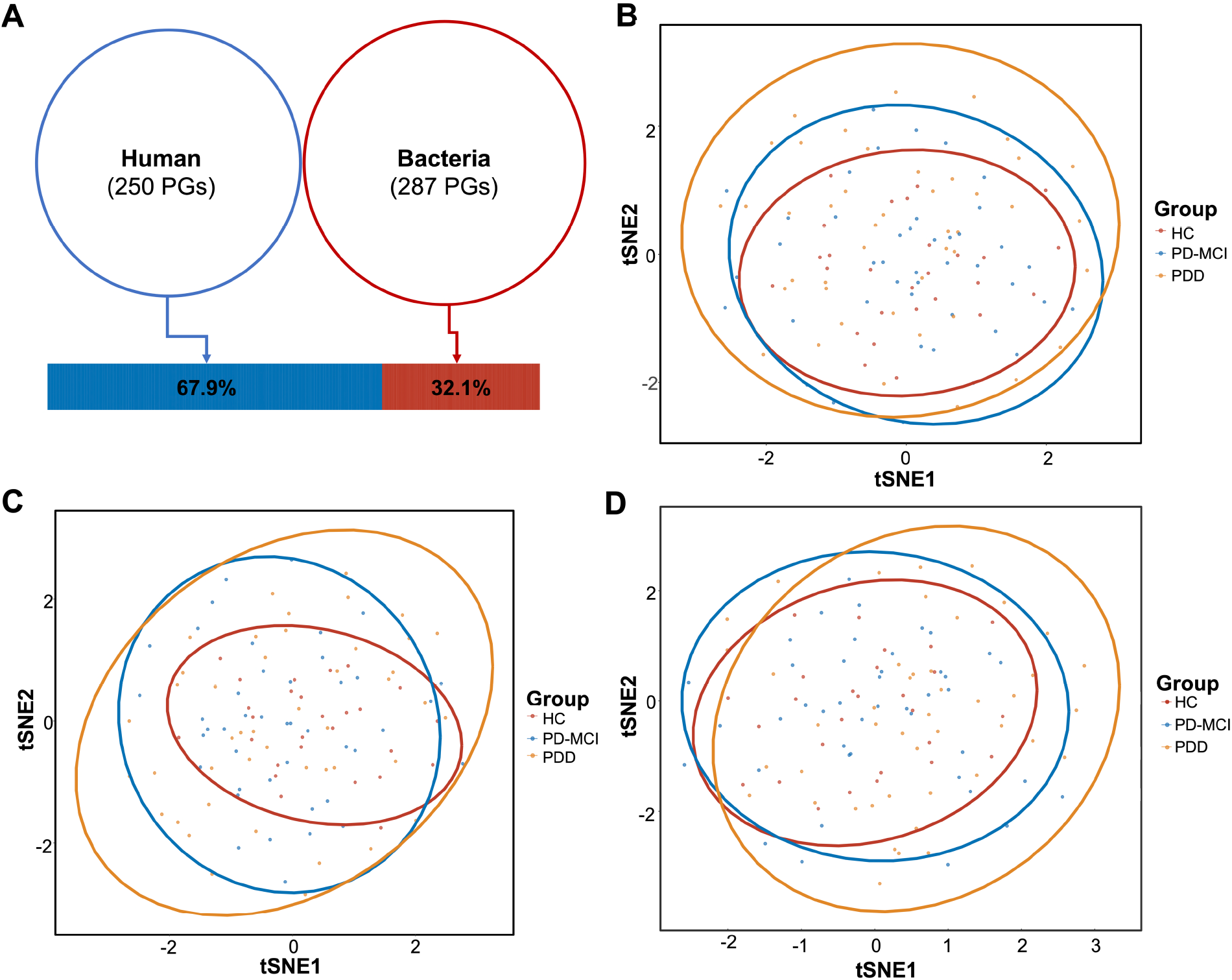
Metaproteome landscape of both human and microbiota protein groups in saliva samples. (A) Distribution of quantified human and microbial protein groups. Venn diagram indicates the numbers of quantified protein groups, while the bar graph shows the total intensity of human or microbial proteins. (B) t-Distributed Stochastic Neighbor Embedding (t-SNE) plot of all proteins quantified in saliva samples (PERMANOVA, R^2^ = 0.026, *p = 0.025*) (C) tSNE plot of bacterial protein groups quantified in saliva samples (PERMANOVA, R^2^ = 0.20, *p* = 0.089). (D) tSNE plot of human protein groups quantified in saliva samples (PERMANOVA, R^2^ =0.21, p = 0.106). Beta diversity comparisons were performed using Aitchison distance. Ellipses represent an 95% confidence level. Color is indicative of the study group.

In order to assess changes in the functional profile of samples, we annotated protein groups using Prophane and applied differential abundance tests both at OG and functional category levels. We have obtained 371 OGs from 24 functional categories and 25 differentially abundant OGs between study groups. At the OG level analysis, functions related to the replication, recombination, and repair (5 OGs in category L), cytoskeleton (4 OGs in category Z), energy production and conversion (3 OGs in category C), translation, ribosomal structure, and biogenesis (2 OGs in category J) were among the most significantly different categories between study groups. 20 of differentially abundant OGs were significantly increased with the progression of CI while 5 OGs showed an increase from HC to MCI and a decrease from MCI to PDD (Figure 5A). The differential abundance results for all OGs and functional categories are shown in Supplementary Figure 5 and Supplementary Figure 6, respectively.

**Figure 5.**
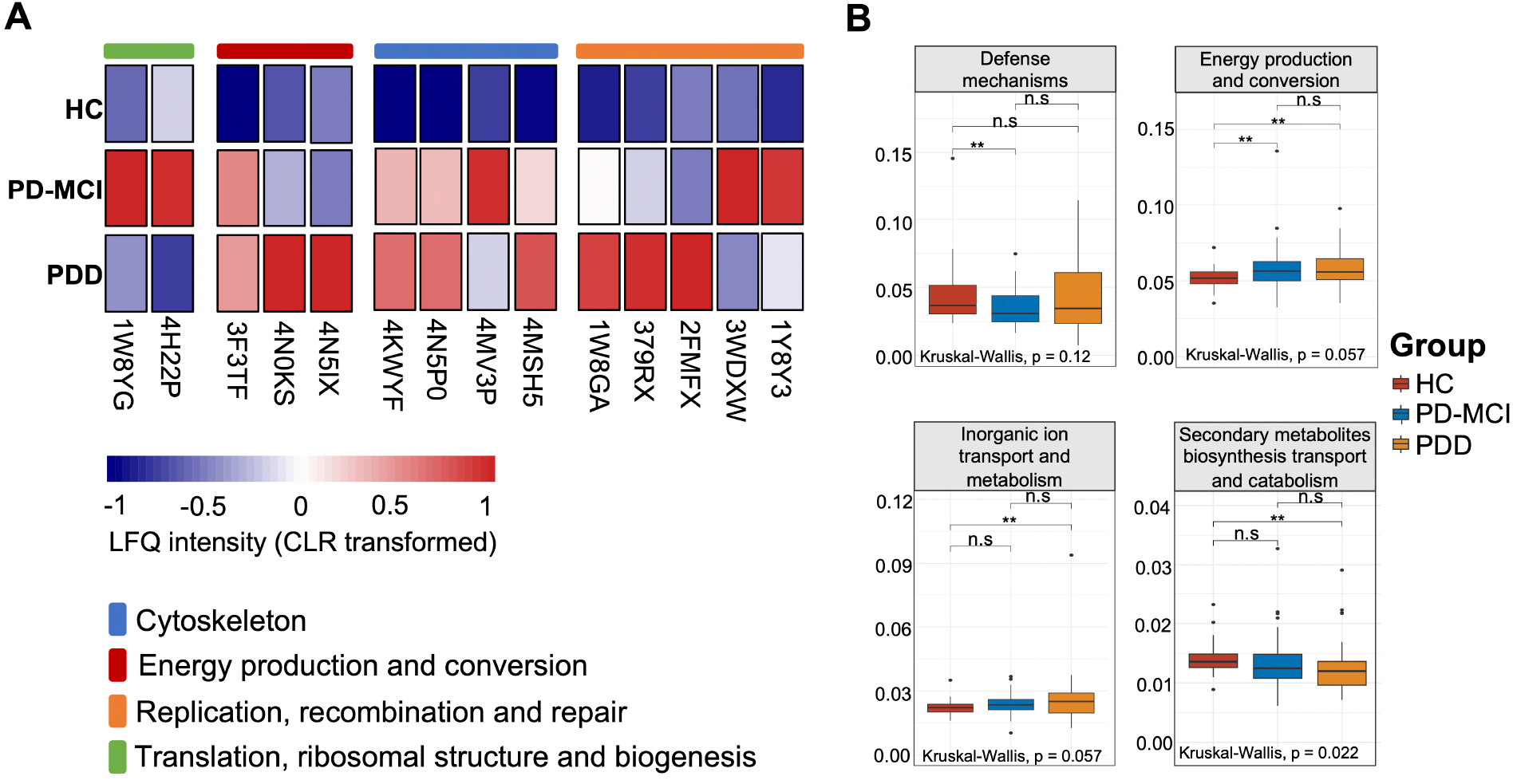
Functional categories of differentially abundant proteins in the saliva samples. (A) Heatmap of differentially abundant OGs between the study groups. Representative OG categories are shown, and the colors indicate the average label-free quantification (LFQ) intensity for each study group. Each row corresponds to an OG with the OG id. CLR: Centered log ratio. (B) LFQ intensity of functional category V (defense mechanism), category C (energy production and conversion), category P (inorganic ion transport and metabolism) and category Q (secondary metabolites biosynthesis transport and catabolism) in saliva samples across the study groups. Median estimates compared across study groups using the Kruskal-Wallis test. Boxes represent the interquartile range, lines indicate medians, and whiskers indicate the range. n.s: not significant, *p < 0.1, **p < 0.05.

At the functional category level, energy production and conversion and inorganic ion transport and metabolism categories were significantly increased with the progression of CI while defense mechanisms and secondary metabolites biosynthesis, transport and catabolism displayed a decreased abundance (Figure 5B).

### 3.3. Multi-omics factor analysis (MOFA)

We applied MOFA to integrate amplicon sequencing and metaproteome results. The fitted model explained 30.1% and 34.4% of the variance in 16S rRNA sequencing and metaproteomics datasets, respectively (Figure 6A) with latent 15 factors (Figure 6B). We examined potential correlations between factors and covariates. We observed that only two of these factors (Factor 2 and Factor 9) were significantly associated with CI in PD and included both bacterial genera and protein features (Figure 6C). Factor 9 has been found to be correlated with Age covariate along with CI stages (Supplementary Figure 7). Thus, we focused on Factor 2 because it was the only factor associated with the progression of CI (R^2^ = 0.064, *p* = 0.003) and identified the contributing features of this factor. Factor 2 values positively correlated with the progression of CI (Figure 6D). Amplicon sequencing based microbiome component of the Factor 2 revealed a decreased abundance of the *Neisseria, Alloprevotella, TM7x, Unclassified Absconditabacteriales* and *Fusobacterium* and increased abundance of *Ligilactobacillus, Lactobacillus, Rothia, Corynebacterium* and *Gemella* in association with the progression of CI in PD. Metaproteome component of Factor 2 showed increased abundance of proteins associated with translation, ribosomal structure and biogenesis (category J), intracellular trafficking, secretion, and vesicular transport (category U), replication, recombination and repair (category L) and unknown function (category S) and a decreased abundance of proteins associated with carbohydrate transport and metabolism (category G) and defense mechanisms (category V).

**Figure 6.**
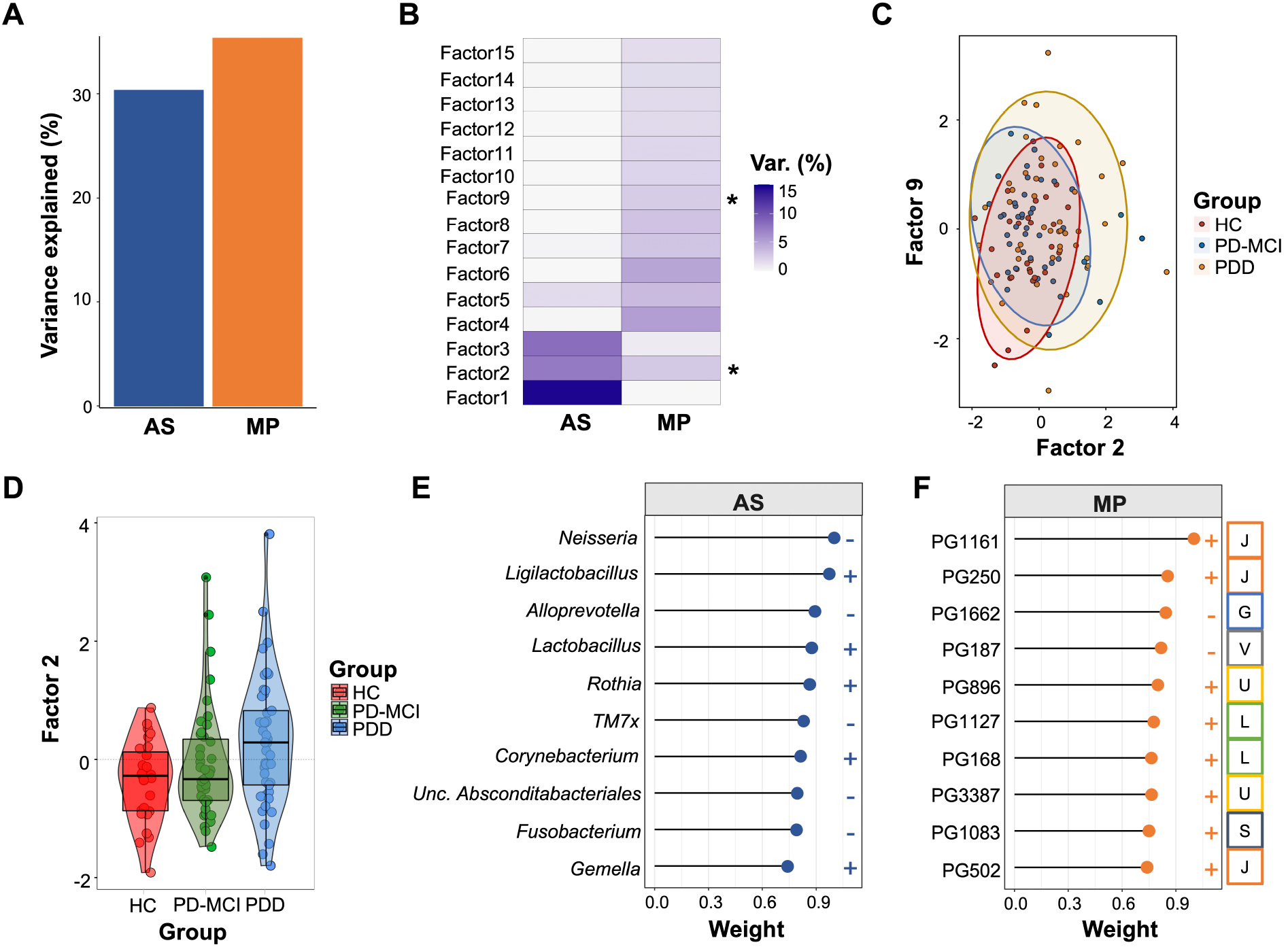
Multi-omics factor analysis (MOFA) of metaproteogenomics dataset. (A) Fraction of total variance explained by type of measurement (view). (B) Fraction of total variance explained by latent factors (LFs) 1–15. (C) Scatter plot of latent Factor 2 (x axis) and latent Factor 9 (y axis). Dots and ellipses are colored according to their group assignment. (D) Box plot of latent Factor 2 values grouped and colored by CI status. (E) Lollipop plot shows top ranking bacterial genera in latent Factor 2. (F) Lollipop plot shows top ranking salivary PGs in latent Factor 2. Human protein groups are colored orange, bacteria protein groups are colored blue.

After examining the contribution of individual features, we selected features with absolute normalized loadings > 0.8 within Factor 2 derived from the MOFA model. This approach selected 7 bacterial genera and 4 proteins with highest weights (Figure 6E). We next decided to generate a machine learning model to assess the ability of these features to predict CI stage of study participants.

### 3.4 Machine learning model

We have generated random forest machine learning models based on 11 features with highest weights within Factor 2 identified by MOFA. We established predictive models based only on bacterial genera (7 features), protein groups (4 features) and a combination of bacterial genera and protein groups (11 features).

Machine learning model based only on bacterial genera showed moderate predictive accuracy for all study groups (HC, AUC = 0.73; PD-MCI, AUC = 0.74; PDD, AUC = 0.72) (Figure 7A) while the predictive model based only on protein groups showed weakest predictive accuracy (HC, AUC = 0.65; PD-MCI, AUC = 0.64; PDD, AUC = 0.57) (Figure 7B). Integrated analysis of the bacterial genera and the protein groups had highest predictive performance (HC, AUC = 0.86; PD-MCI, AUC = 0.81; PDD, AUC = 0.74) (Figure 7C) which indicates that integrating features from different data modalities leads to an increase in the predictive performance. We determined and plotted mean decrease in Gini values which showed PG1662 and *Neisseria* as the strongest predictive features for CI stage in PD (Figure 7D).

**Figure 7.**
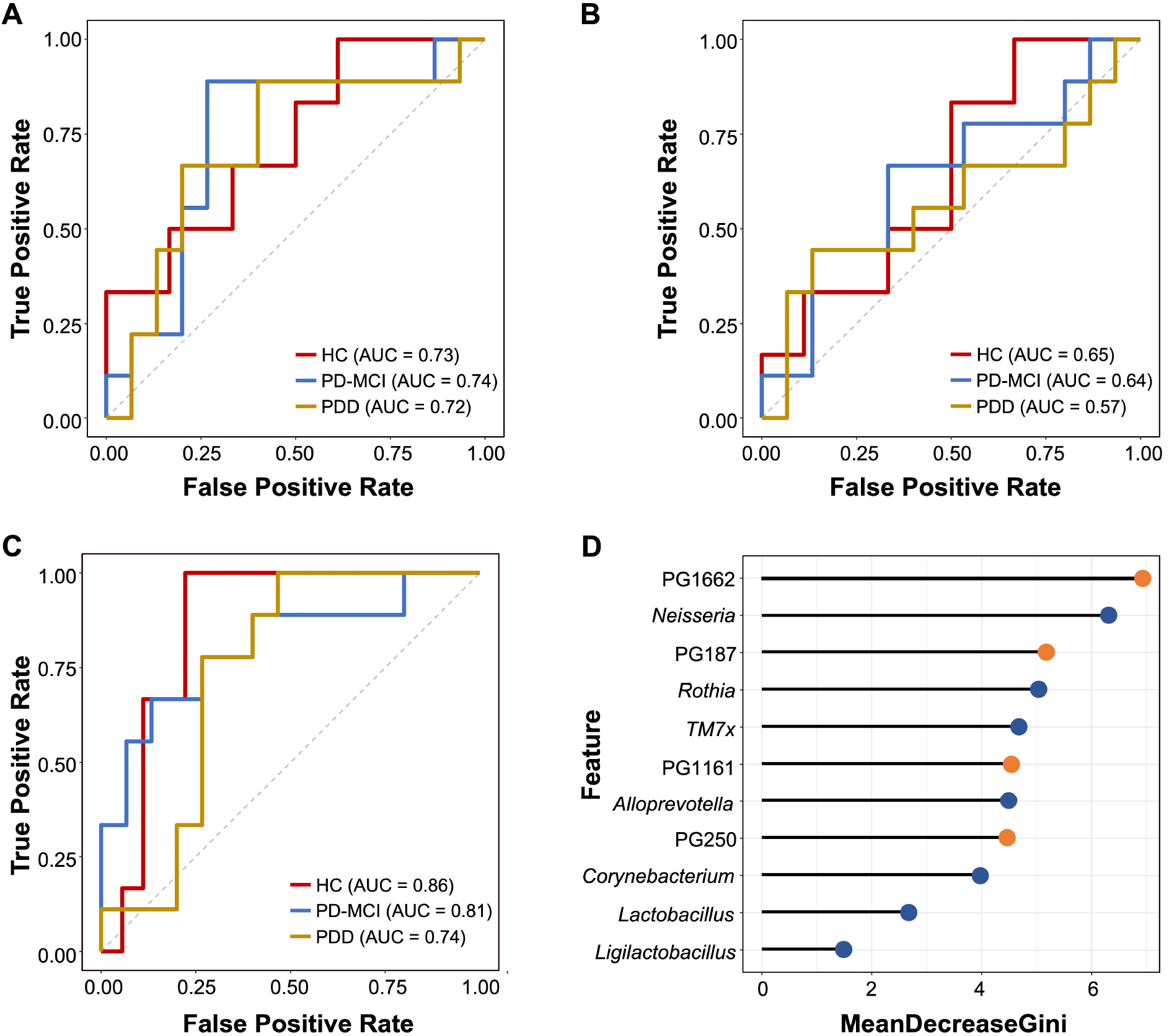
Random forest model of MOFA selected features from bacterial genera and metaproteomics results. (A) Receiver operating characteristic (ROC) curves for MOFA selected bacterial genera (weight >0.8) to classify participants based on CI status. AUC, area under the curve. Line color is indicative of the study group. (B) Receiver operating characteristic (ROC) curves for MOFA selected proteins (weight >0.8) to classify participants based on CI status. (C) Receiver operating characteristic (ROC) curves for MOFA selected both bacterial genera (weight >0.8) and proteins (weight >0.8) to classify participants based on CI status. (D) Mean decrease in Gini values for 11 features including bacterial genera and proteins used in the random forest model. Dots are colored blue and orange for PG and bacterial genera, respectively.

To strengthen our ML analysis, we also used AutoGluon for prediction of CI stages by MOFA selected features (Supplementary Figure 8). AutoGluon results showed the weakest predictive performance when only 4 PG features were used as input for the model (HC-MCI, AUC=0.53; HC-PDD, AUC=0.54; MCI-PDD, AUC=0.67), in consistent with RF model results. The predictive performances of 7 bacterial genera were better than the predictive performance of 4 PGs (HC-MCI, AUC=0.70; HC-PDD, AUC=0.72; MCI-PDD, AUC=0.58) while using all MOFA selected features increased the predictive performance only in differentiating PDD and MCI groups (HC-MCI, AUC=0.70; HC-PDD, AUC=0.72; MCI-PDD, AUC=0.7).

## 4. Discussion

In this study, we characterized the compositional dynamics of saliva of PD patients across a continuum of CI (PD-MCI and PDD) as compared with HC using both 16S rRNA gene amplicon sequencing and metaproteomics profiling. We identified the discriminatory key signatures in saliva composition related to CI stages of PD by applying an integrative metaproteogenomics approach. We demonstrated that a shift in salivary microbiome and protein translation machinery and defense mechanism related changes in human proteome is associated with the CI progression in PD. In addition, we showed that fusion of these features yields a higher predictive performance for classification of CI stages in PD. To our knowledge, this is the first study reporting integrative analysis of saliva metagenomics and proteomics in association with cognitive decline in PD.

Both metaproteome and microbiota profiles indicated *Streptococcus, Prevotella, Veillonella, Fusobacterium* and *Neisseria* as the most abundant bacterial genera in the study cohort which is in agreement with a previous study comparing salivary microbiota of healthy controls and PD patients^15^. The amplicon sequencing detected much more bacterial genera, families, and phyla than metaproteomics profiling. This is not surprising because only a subset of bacterial genera (as described in the Methods) were used to generate a custom-based reference protein database to identify proteins.

Although alpha diversity in amplicon-based microbiota showed no significant differences between study groups beta diversity comparisons of saliva samples significantly differentiated CI stages, suggesting salivary bacterial community restructures by the progression of CI. Results of four differential abundance methods showed in consensus a significant decrease of *Neisseria* genus with the progression of CI. Previous studies have already shown a decreased abundance of *Neisseria* and *Neisseriaceae* in PD oral swab samples^39,40^. Here, we further showed the continuous decrease of *Neisseria* with the progression of CI in PD.

Metaproteome profile of saliva samples was composed of a balanced number of human and bacterial protein groups. On the other hand, human proteins constituted much higher percentage of total protein intensities measured in the saliva samples, as observed in previous studies^38^. Differential abundance analyses of metaproteome profiles determined marked functional alterations associated with CI. Particularly, the functions related to cytoskeleton and translation, ribosomal structure and biogenesis, defense mechanisms and energy production and conversion were among the most significantly altered functions between study groups.

Big data from multi-omics studies cause paradigm shift in modern medicine. Because data from single modality studies often fail to capture desired predictive performance of the disease progression, we selected the multimodal features with the highest weights to test whether we can predict CI stages of PD patients by ML models. The integrated ML model included 7 bacterial genera namely, *Neisseria, Lactobacillus, Rothia, Ligilactobacillus, Alloprevotella, TM7x* and *Corynebacteirum* and 4 protein groups which were annotated as 40S ribosomal protein SA (RPSA), 40S ribosomal protein S15 (RPS15), pyruvate, phosphate dikinase (PPDK) and bactericidal permeability-increasing protein (BPI). We determined that the integrated ML model based on this combinatorial marker panel shows better predictive performance than the ML models based on features from single modality. Together, these alterations point to a multi-layer disturbance of salivary composition in association with CI in PD as discussed below.

The taxa *Neisseria* deserves a special attention among others as it is implicated in prevention of oral diseases due to its beneficial abilities such as metabolizing low-pH products into weak acids. On the other hand, consistently reported high abundance of *Lactobacillaceae* members in the oral cavity of PD patients may have negative effects due to their ability to reduce secretion of neuroprotective hormones such as ghrelin^41^. The decrease in *Neisseria* accompanied with the increased abundance of *Lactobacillaceae* family has been also reported in a recent study on PD^16^. Our results suggest a continuity for this compositional shift across CI spectrum in PD. Moreover, we have detected a decrease in PPDK enzyme, as another signature. PPDK is known as one of key enzymes in gluconeogenesis^42^ and related to lactic acid production^43^. Furthermore, we detected an increased abundance of two ribosomal proteins, RPS15 and RPSA. Both of these ribosomal proteins play a central role in ribosome biogenesis and protein translation. RPS15 has been identified as a pathogenic substrate of LRRK2, mutations of which are the most common genetic etiology of PD^44^ while RPSA has been shown to increase in the presence of fibrillar α-syn^45^. The association of these ribosomal proteins with the progression of CI contributes to the increasing accumulating evidence on the role of deregulated protein translation machinery in PD. BPI, identified as another signature protein group, is involved in the defense of host against bacterial pathogens and considered as a microbial translocation markers^46^. Elevated serum endotoxin levels have been previously reported in PD patients, which indicate greater bacterial translocation, particularly for subgroup with high risk for early dementia^47^. Thus, our observation of decreased BPI abundance in saliva suggests a potential LPS-BPI imbalance in PD patients which worsens with the CI progression.

In conclusion, our study presents a comprehensive overview of significant key changes in saliva composition that parallel the progression of CI in PD and suggests potential non-invasive biomarker candidates for predicting CI in PD. To our knowledge, this is the first study based on the novel approach by integrating amplicon sequencing and metaproteomics for characterization of salivary composition of PD patients and investigating its association with CI stages in PD. Metaproteogenomics can provide useful non-invasive tools for better monitoring and classification of CI stages.

## Data Availability

The 16S rRNA gene amplicon sequencing data produced in this study have been deposited in the NCBI Sequence Read Archive database under accession no. PRJNA913682

https://www.ncbi.nlm.nih.gov/sra/PRJNA913682

## Acknowledgements

This study was supported by a grant to Suleyman Yildirim from the Scientific and Technological Research Council of Turkey (TUBITAK) (grant no. 315S301). Muzaffer Arıkan was a recipient of FEMS Research and Training Grant. The authors declare no conflicts of interest.

## Author contributions

Conception and Design, S.Y., L.H., and M.Ö; Sample Collection and Processing, M.A., T.K.D., Z.Y., N.H.Y., A.S., L.H., N.D.K.; Data Analysis, M.A., OUN; Data Interpretation, M.A., S.Y., and T.M.; Manuscript Writing – Original Draft, M.A., S.Y.; Review & Editing, S.Y., M.Ö., and T.M. All authors read and approved the final manuscript.

## References

1. Vos, T. et al. Global, regional, and national incidence, prevalence, and years lived with disability for 301 acute and chronic diseases and injuries in 188 countries, 1990–2013: a systematic analysis for the Global Burden of Disease Study 2013. Lancet 386, 743–800 (2015).

2. Dorsey, E. R. et al. Projected number of people with Parkinson disease in the most populous nations, 2005 through 2030. Neurology 68, 384–6 (2007).

3. Kalia, L. V & Lang, A. E. Parkinson’s disease. Lancet 386, 896–912 (2015).

4. O’Callaghan, C. & Lewis, S. J. G. Cognition in Parkinson’s Disease. in International Review of Neurobiology vol. 133 557–583 (Elsevier Inc., 2017).

5. Aarsland, D. et al. Parkinson disease-associated cognitive impairment. Nat. Rev. Dis. Prim. 7, 47 (2021).

6. Aro, K., Wei, F., Wong, D. T. & Tu, M. Saliva Liquid Biopsy for Point-of-Care Applications. Front. Public Heal. 5, (2017).

7. Figura, M. & Friedman, A. In search of Parkinson’s disease biomarkers - is the answer in our mouths? A systematic review of the literature on salivary biomarkers of Parkinson’s disease. Neurol. Neurochir. Pol. 54, 14–20 (2020).

8. Vivacqua, G. et al. Abnormal Salivary Total and Oligomeric Alpha-Synuclein in Parkinson’s Disease. PLoS One 11, e0151156 (2016).

9. Kang, W. et al. Salivary total α-synuclein, oligomeric α-synuclein and SNCA variants in Parkinson’s disease patients. Sci. Rep. 6, 28143 (2016).

10. Vivacqua, G. et al. Salivary alpha-synuclein in the diagnosis of Parkinson’s disease and Progressive Supranuclear Palsy. Parkinsonism Relat. Disord. 63, 143–148 (2019).

11. Adler, C. H. & Beach, T. G. Neuropathological basis of nonmotor manifestations of Parkinson’s disease. Mov. Disord. 31, 1114–1119 (2016).

12. Mu, L. et al. Alpha-Synuclein Pathology in Sensory Nerve Terminals of the Upper Aerodigestive Tract of Parkinson’s Disease Patients. Dysphagia 30, 404–417 (2015).

13. Wang, P. et al. Six-Year Follow-Up of Dysphagia in Patients with Parkinson’s Disease. Dysphagia 37, 1271–1278 (2022).

14. van Wamelen, D. J. et al. Drooling in Parkinson’s Disease: Prevalence and Progression from the Non-motor International Longitudinal Study. Dysphagia 35, 955–961 (2020).

15. Fleury, V. et al. Oral Dysbiosis and Inflammation in Parkinson’s Disease. J. Parkinsons. Dis. 11, 619–631 (2021).

16. Rozas, N. S., Tribble, G. D. & Jeter, C. B. Oral Factors That Impact the Oral Microbiota in Parkinson’s Disease. Microorganisms 9, 1616 (2021).

17. Litvan, I. et al. Diagnostic criteria for mild cognitive impairment in Parkinson’s disease: Movement Disorder Society Task Force guidelines. Mov. Disord. 27, 349–56 (2012).

18. Arikan, M. et al. Axillary Microbiota Is Associated with Cognitive Impairment in Parkinson’s Disease Patients. Microbiol. Spectr. 10, e0235821 (2022).

19. Beker, M. C. et al. Interaction of melatonin and Bmal1 in the regulation of PI3K/AKT pathway components and cellular survival. Sci. Rep. 9, 19082 (2019).

20. Weber, N. et al. Nephele: a cloud platform for simplified, standardized and reproducible microbiome data analysis. Bioinformatics 34, 1411–1413 (2018).

21. Quast, C. et al. The SILVA ribosomal RNA gene database project: Improved data processing and web-based tools. Nucleic Acids Res. 41, 590–596 (2013).

22. Davis, N. M., Proctor, D. M., Holmes, S. P., Relman, D. A. & Callahan, B. J. Simple statistical identification and removal of contaminant sequences in marker-gene and metagenomics data. Microbiome 6, 226 (2018).

23. Bolyen, E. et al. Reproducible, interactive, scalable and extensible microbiome data science using QIIME 2. Nat. Biotechnol. 37, 852–857 (2019).

24. McMurdie, P. J. & Holmes, S. phyloseq: an R package for reproducible interactive analysis and graphics of microbiome census data. PLoS One 8, e61217 (2013).

25. Mallick, H. et al. Multivariable association discovery in population-scale meta-omics studies. PLoS Comput. Biol. 17, e1009442 (2021).

26. Segata, N. et al. Metagenomic biomarker discovery and explanation. Genome Biol. 12, R60 (2011).

27. Lin, H. & Peddada, S. Das. Analysis of compositions of microbiomes with bias correction. Nat. Commun. 11, 3514 (2020).

28. Wickham, H. ggplot2. vol. 10 (Springer New York, 2009).

29. Zhu, J. et al. Over 50,000 Metagenomically Assembled Draft Genomes for the Human Oral Microbiome Reveal New Taxa. Genomics. Proteomics Bioinformatics (2021) doi:10.1016/j.gpb.2021.05.001.

30. UniProt Consortium. UniProt: a worldwide hub of protein knowledge. Nucleic Acids Res. 47, D506–D515 (2019).

31. Ma, S. et al. Population structure discovery in meta-analyzed microbial communities and inflammatory bowel disease using MMUPHin. Genome Biol. 23, 208 (2022).

32. Schneider, T. et al. Structure and function of the symbiosis partners of the lung lichen (Lobaria pulmonaria L. Hoffm.) analyzed by metaproteomics. Proteomics 11, 2752–2756 (2011).

33. Huerta-Cepas, J. et al. eggNOG 5.0: a hierarchical, functionally and phylogenetically annotated orthology resource based on 5090 organisms and 2502 viruses. Nucleic Acids Res. 47, D309–D314 (2019).

34. Argelaguet, R. et al. Multi-Omics Factor Analysis—a framework for unsupervised integration of multi-omics data sets. Mol. Syst. Biol. 14, 1–13 (2018).

35. Argelaguet, R. et al. MOFA+: a statistical framework for comprehensive integration of multi-modal single-cell data. Genome Biol. 21, 111 (2020).

36. Liu, C., Cui, Y., Li, X. & Yao, M. microeco : an R package for data mining in microbial community ecology. FEMS Microbiol. Ecol. 97, 1–9 (2021).

37. Nearing, J. T. et al. Microbiome differential abundance methods produce different results across 38 datasets. Nat. Commun. 13, 1–10 (2022).

38. Granato, D. C. et al. Meta-omics analysis indicates the saliva microbiome and its proteins associated with the prognosis of oral cancer patients. Biochim. Biophys. Acta - Proteins Proteomics 1869, 140659 (2021).

39. Pereira, P. A. B. et al. Oral and nasal microbiota in Parkinson’s disease. Parkinsonism Relat. Disord. 38, 61–67 (2017).

40. Li, Z. et al. Oral, Nasal, and Gut Microbiota in Parkinson’s Disease. Neuroscience 480, 65–78 (2022).

41. Mihaila, D. et al. The oral microbiome of early stage Parkinson’s disease and its relationship with functional measures of motor and non-motor function. PLoS One 14, e0218252 (2019).

42. Wu, C., Dunaway-Mariano, D. & Mariano, P. S. Design, Synthesis, and Evaluation of Inhibitors of Pyruvate Phosphate Dikinase. J. Org. Chem. 78, 1910–1922 (2013).

43. Yamada, T. & Carlsson, J. Regulation of lactate dehydrogenase and change of fermentation products in streptococci. J. Bacteriol. 124, 55–61 (1975).

44. Martin, I. et al. Ribosomal protein s15 phosphorylation mediates LRRK2 neurodegeneration in Parkinson’s disease. Cell 157, 472–485 (2014).

45. Pieri, L. et al. Cellular response of human neuroblastoma cells to α-synuclein fibrils, the main constituent of Lewy bodies. Biochim. Biophys. Acta - Gen. Subj. 1860, 8–19 (2016).

46. Krasity, B. C., Troll, J. V., Weiss, J. P. & McFall-Ngai, M. J. LBP/BPI proteins and their relatives: Conservation over evolution and roles in mutualism. Biochem. Soc. Trans. 39, 1039–1044 (2011).

47. Wijeyekoon, R. S. et al. Peripheral innate immune and bacterial signals relate to clinical heterogeneity in Parkinson’s disease. Brain. Behav. Immun. 87, 473–488 (2020).

